# Anti-Neuraminidase Antibodies Reduce the Susceptibility to and Infectivity of Influenza A/H3N2 Virus

**DOI:** 10.1101/2024.06.14.24308936

**Authors:** Gregory Hoy, Thomas Cortier, Hannah E. Maier, Guillermina Kuan, Roger Lopez, Nery Sanchez, Sergio Ojeda, Miguel Plazaola, Daniel Stadlbauer, Abigail Shotwell, Angel Balmaseda, Florian Krammer, Simon Cauchemez, Aubree Gordon

**Author notes:** Denotes co-senior authors (Simon Cauchemez, Aubree Gordon).

## Abstract

Immune responses against neuraminidase (NA) are of great interest for developing more robust influenza vaccines, but the role of anti-NA antibodies on influenza infectivity has not been established. We conducted household transmission studies in Managua, Nicaragua to examine the impact of anti-NA antibodies on influenza A/H3N2 susceptibility and infectivity. Analyzing these data with mathematical models capturing household transmission dynamics and their drivers, we estimated that having higher preexisting antibody levels against the hemagglutinin (HA) head, HA stalk, and NA was associated with reduced susceptibility to infection (relative susceptibility 0.67, 95% Credible Interval [CrI] 0.50-0.92 for HA head; 0.59, 95% CrI 0.42-0.82 for HA stalk; and 0.56, 95% CrI 0.40-0.77 for NA). Only anti-NA antibodies were associated with reduced infectivity (relative infectivity 0.36, 95% CrI 0.23-0.55). These benefits from anti-NA immunity were observed even among individuals with preexisting anti-HA immunity. These results suggest that influenza vaccines designed to elicit NA immunity in addition to hemagglutinin immunity may not only contribute to protection against infection but reduce infectivity of vaccinated individuals upon infection.

## Introduction

Influenza virus infection remains an important cause of human disease burden, with upwards of one billion infections and up to 650,000 deaths occurring globally every year from disease caused by the influenza virus^1^. Vaccination against influenza virus is one of the most effective approaches for reducing the overall morbidity and mortality of seasonal influenza in communities, and improving the effectiveness of influenza vaccines is an important goal^2–5^. There are two important components of transmission; *susceptibility* refers to an individual or group’s propensity to become infected with influenza, assuming adequate exposure. *Infectivity* refers to an individual or group’s propensity to infect others, assuming that they themselves are infected. Conditional on the first person already being infected, the overall risk of transmission from one person to another depends on the infectivity of the first person and the susceptibility of the second. Much of the effort for influenza vaccine improvement has focused on the induction of immune responses that reduce susceptibility to infection or disease, to moderate effectiveness; much less attention has been given to the development of vaccines that reduce individual-level infectivity among the vaccinated; in other words, vaccines that generate an immune response that decreases the infectivity of vaccinees, even if they are not fully protected from infection^3,4,6–8^. Population-level vaccination efforts clearly reduce overall community transmission, likely because vaccinated individuals show reduced susceptibility to infection, which breaks transmission chains^9–11^. However, there is no evidence that current-generation influenza vaccines reduce individual-level infectivity directly, and little is known about if and how immune responses that protect against influenza virus infection affect individual infectivity. The identification of immune responses that both reduce susceptibility to infection and reduce infectivity among vaccinated individuals who are infected would allow for the development of influenza vaccinations that lower overall influenza circulation in communities and that provide additional indirect protection to individuals who are unvaccinated or under vaccinated for influenza, including infants and immunocompromised individuals.

Antibody responses against neuraminidase (NA), an influenza surface glycoprotein, are thought to protect against severe disease caused by the influenza virus, and high pre-infection anti-NA antibody levels have been shown to reduce the overall duration of influenza viral shedding^12–19^. Additionally, studies in animal models have demonstrated a reduction in viral shedding occurring in animals immunized against neuraminidase^20,21^. However, viral shedding does not always consistently correlate with infectivity, and direct transmission reduction of anti-NA immunity has not been demonstrated in human populations^22^. Additionally, the role of the anti-NA response on infectivity, relative to other important immune targets such as hemagglutinin, has not been investigated. This study aims to explore the impact of pre-existing antibody levels against the hemagglutinin (HA) head, HA stalk, and NA on influenza virus A/H3N2 transmission in a household setting, with particular interest in the role of anti-neuraminidase responses in modulating transmission risk.

## Results

### Participant and household characteristics

Over three influenza seasons (2014, 2016, 2017), a total of 171 households (171 index cases and their 664 households contacts) were recruited following the detection of an infected individual (i.e. the index case) and followed up for an average duration of 36.7 days. 148 out of 664 (22.3%) household contacts were infected during the follow-up period. Households were enrolled through identification of an index case at the study clinic (2014, 2016), or pre-enrolled households were activated after detection of influenza virus via polymerase chain reaction (PCR) in a member of an enrolled household (2017). Once activated, index cases and household contacts were tested for influenza virus every 2-3 days using PCR, and serology was done on blood samples collected on the first day of household activation (the initial/acute sample) and 30-45 days after household activation (the final/convalescent sample). More information about the study design and case ascertainment is available in the Online Methods. The number of infections per household ranged from 1 to 10, with an average of 1.87 total infections and 0.87 secondary infections per household. The serial interval (i.e. the average time between index case onset and onset of cases in household contacts) was 3.4 days (SD 2.8 days). A visualization of the intensive monitoring periods by household is presented in Figure 1.

**Figure 1.**
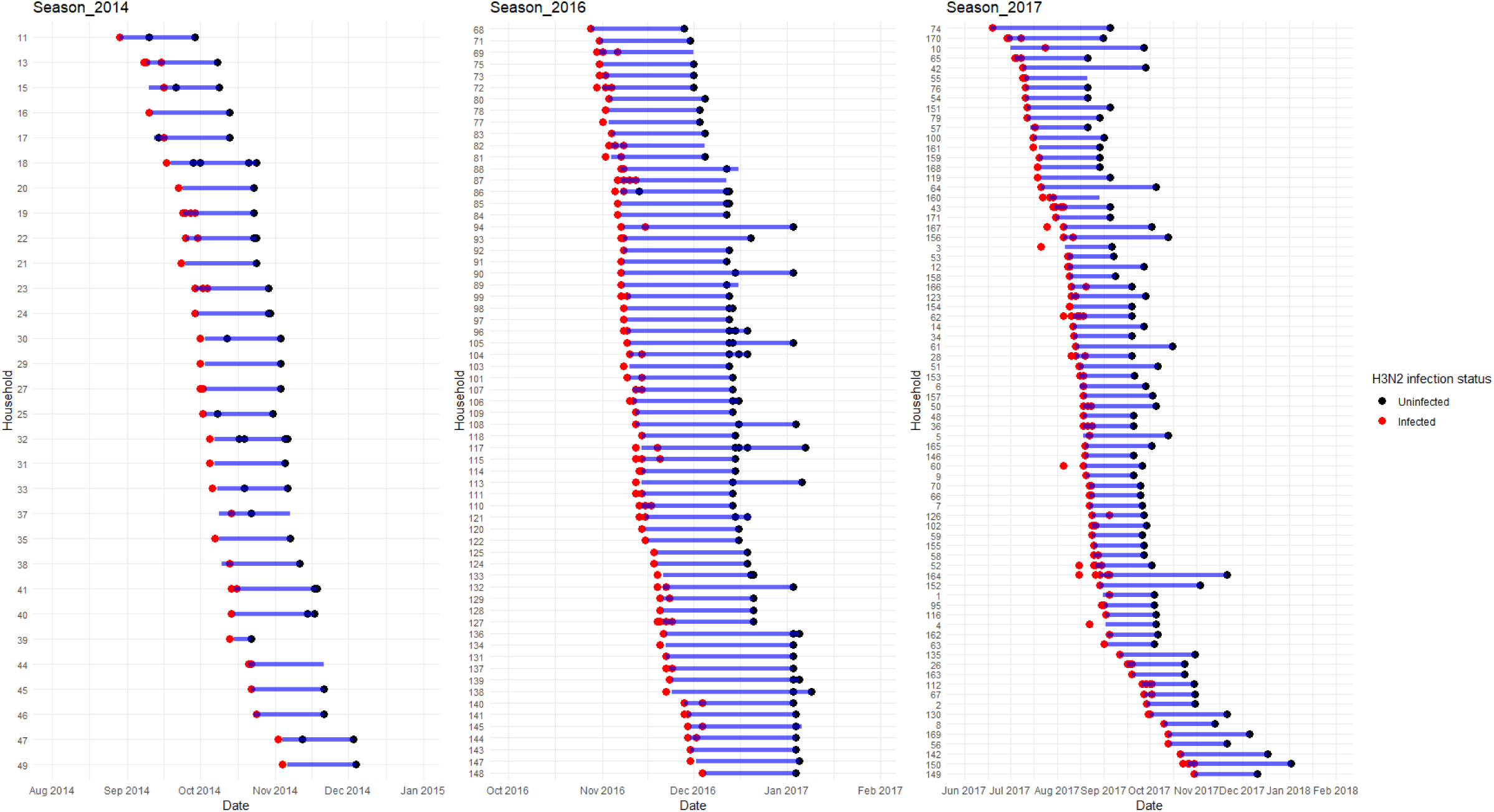
Intensive monitoring periods by household. Infected individuals represented with red dots; uninfected individuals are represented in blue dots. The blue bars represent the duration of the influenza intensive monitoring period within the household.

Less than 10% of individuals had ever been vaccinated against influenza, and only two individuals had been vaccinated for influenza in the 6 months prior to the start of the monitoring period. Positive individuals were younger. Overall, a higher proportion of infected individuals had anti-HA head, HA stalk, and NA pre-existing antibody levels in the lower quartiles when compared to uninfected individuals (Table 1). There was no difference in the pre-existing antibody levels between index cases and secondary cases for hemagglutination inhibition assay titers (HAI) and HA stalk antibody levels; however, index cases had slightly lower anti-NA antibody levels when compared to secondary cases (median AUC 29.7 and 45.4, respectively, p = 0.042) (Figure 2a). The secondary attack rate in households with an index case with low-to-undetectable anti-NA antibodies was 23.4%, compared to 17.1% in households with an index case with higher anti-NA antibodies (p=0.15); there was no difference in secondary attack rates in households by index case anti-HA head or anti-HA stalk antibodies (Figure 2b).

**Figure 2.**
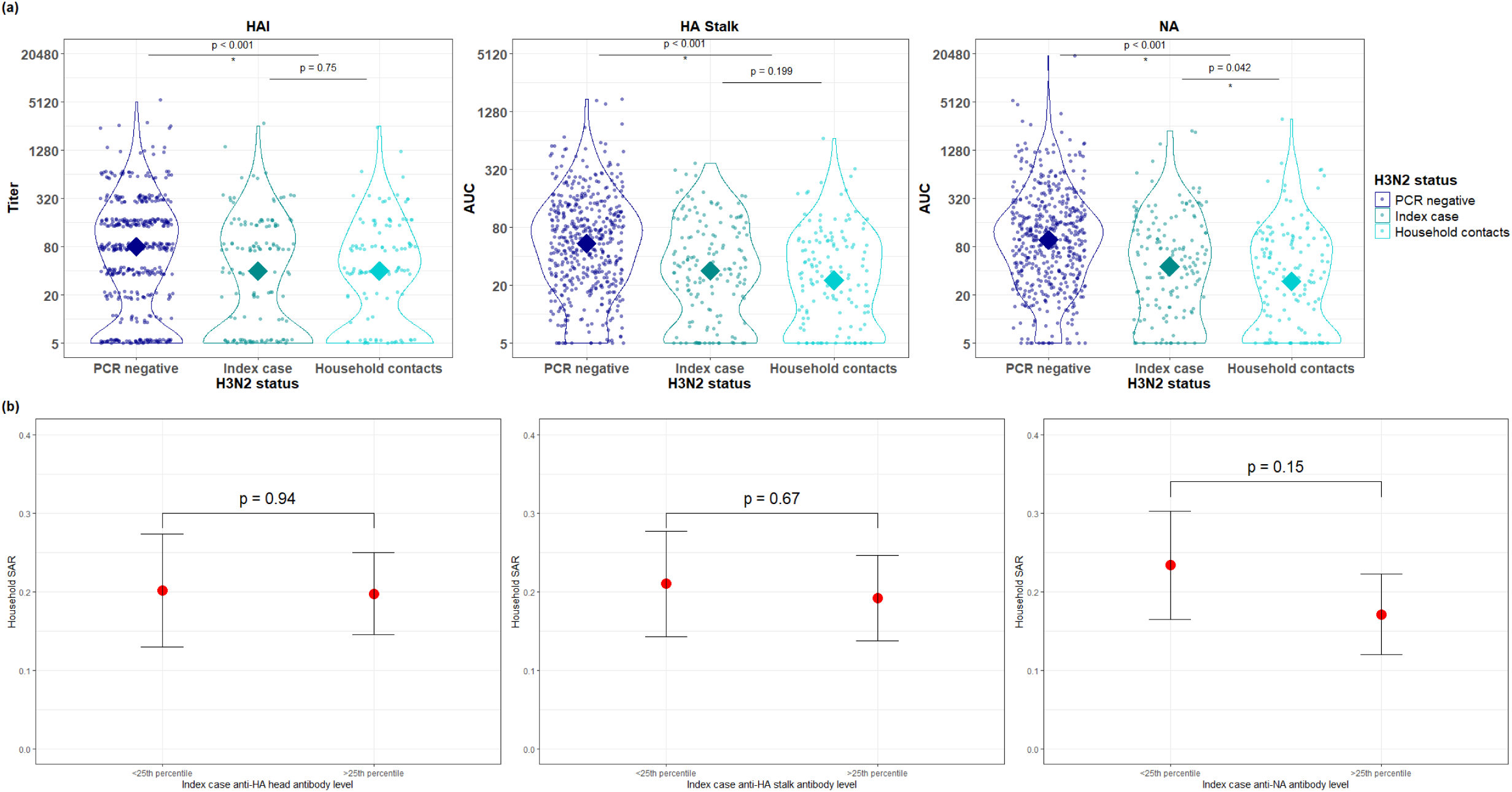
Antibody levels by PCR status and household SAR. (a) Distribution of pre-infection antibody titers by PCR-negative individuals, probable index cases, and probable secondary/tertiary cases (chevrons representing mean antibody level). (b) Mean and 95% confidence intervals for the secondary attack rates by household, stratified by the antibody levels of the household index case. P-values are calculated from generalized estimating equations (GEEs) with weighting by household size.

**Table 1.**
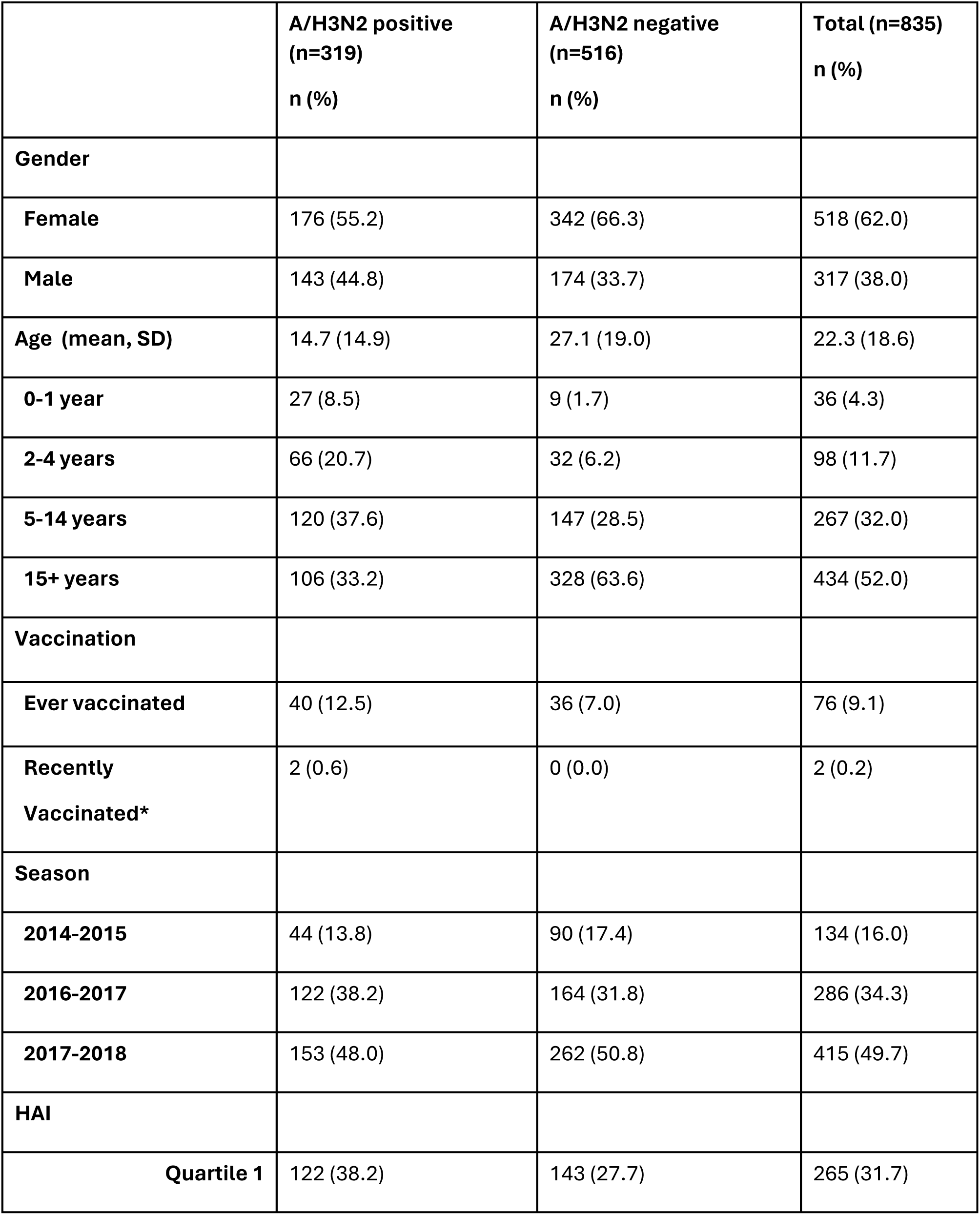
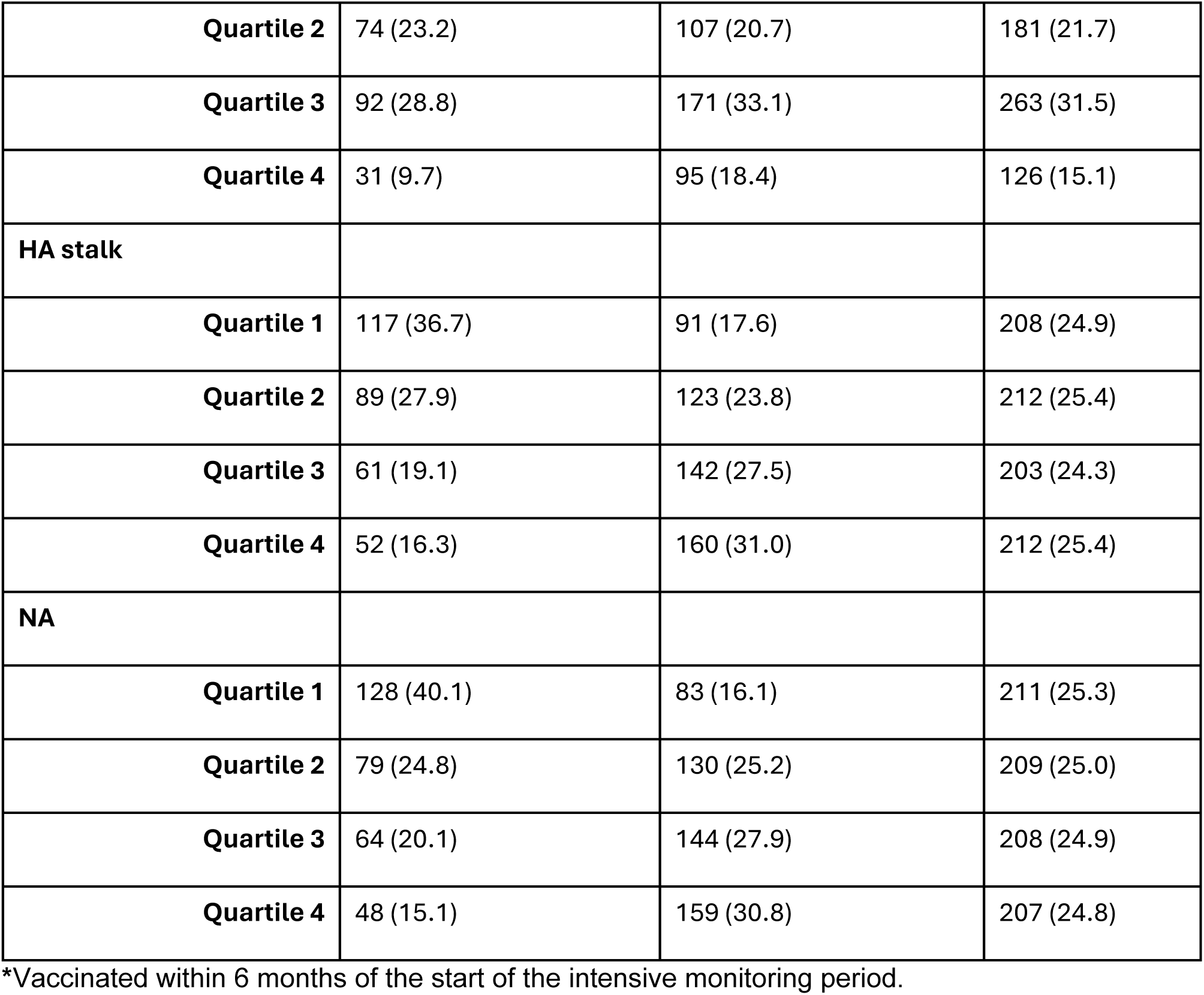
Description of Study Population.

### Effect of pre-existing antibody levels on susceptibility and infectivity

A mathematical model, calibrated to the data with Bayesian data augmentation methods, was used to reconstruct the unobserved chains of transmission accounting for the possibility of community (i.e. household member infected outside the household) and tertiary (i.e. household member infected by another household member who is not the index case) infections, estimate household transmission rates and determine factors affecting individual relative susceptibility and infectivity (see Online Methods)^23^.

Compared to adults 15+ years of age, children aged 0-14 years had higher relative susceptibility (relative susceptibility 1.63, 95% CrI 1.22-2.18). High initial antibody levels against the HA head (0.67, 95% CrI 0.50-0.92), HA stalk (0.59, 95% CrI 0.42-0.82), and NA (0.56, 95% CrI 0.40-0.77) were associated with reduced susceptibility to influenza A/H3N2 virus infection (Figure 3a). In infected A/H3N2 individuals, high initial antibody levels against NA were associated with reduced infectivity (relative infectivity 0.36, 95% CrI 0.23-0.55). In contrast, high initial antibody levels against the HA head and the HA stalk were not associated with reduced infectivity (1.33, 95% CrI 0.90-1.90 for the HA head, 1.08, 95% CrI 0.75-1.59 for the HA stalk)(Figure 3b). The probability of infection from the community was estimated to be 5.8% per month (95% CrI 2.7%-10.8%).

**Figure 3.**
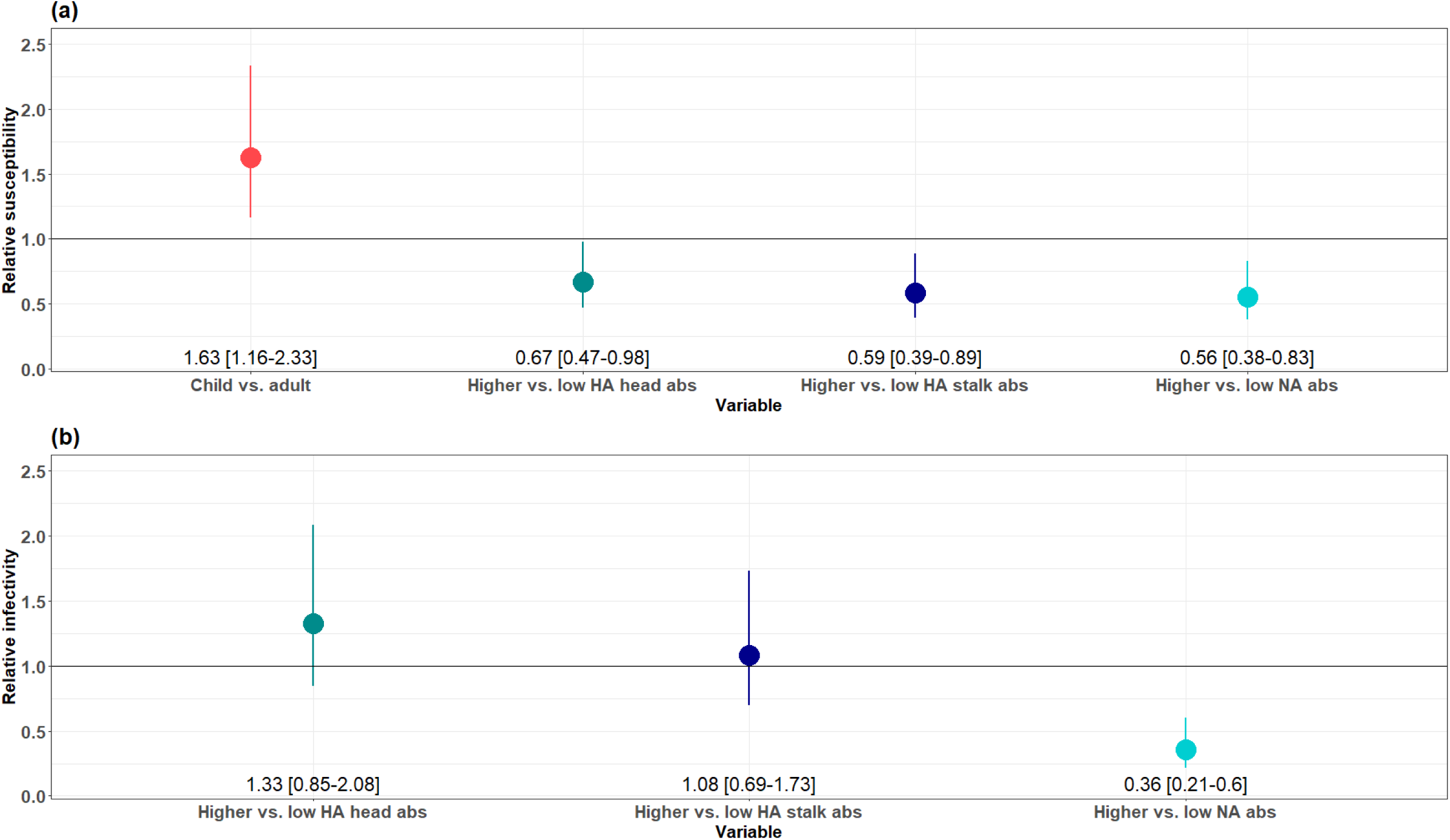
Relative susceptibility and infectivity of A/H3N2 by age and antibody levels. (a) Point estimate and 95% credible interval for relative susceptibility to A/H3N2 as a function of age and high versus low initial antibody levels against the HA head, HA stalk, and NA. (b)) Point estimate and 95% credible interval for relative infectivity of A/H3N2 as a function of high versus low initial antibody levels against the HA head, HA stalk, and NA.

### Necessity of robust antibody responses to reduce susceptibility

Because antibodies against all three antigens were associated with reduced susceptibility, we next explored whether high antibody levels against a single antigen could significantly impact susceptibility and infectivity, or whether a combination of immune responses was needed. We therefore compared the A/H3N2 susceptibility and infectivity of individuals who had high antibody levels for zero antibody measures, one antibody measure, two antibody measures where none are against NA, and two or more antibody measures where one is against NA. This model allows us to test the hypothesis of an additive protective effect of anti-HA head, anti-HA stalk and anti-NA antibodies made in our baseline model, as well as any dose-response pattern observed in the relationship between antibody levels and susceptibility/infectivity. Furthermore, by splitting the high-responder categories by those with anti-NA antibodies and those with low-to-undetectable anti-NA antibodies, we are able to further test whether anti-NA immunity is uniquely associated with reduced infectivity in influenza virus A/H3N2. Individuals with higher antibody levels for one measure did not see their susceptibility or infectivity modified compared to individuals with low antibody levels for all measures (relative susceptibility 0.83, 95% CrI 0.49-1.38; relative infectivity 0.95, 95% CrI 0.56-1.52), nor did individuals with higher antibody levels for both the anti-HA measures (relative susceptibility 0.62, 95% CrI 0.34-1.10; relative infectivity 1.17, 95% CrI 0.62-2.03). However, individuals with high antibody levels for two or more measures, one of which is a response against NA, demonstrate reduced susceptibility to and infectivity of influenza virus A/H3N2 (relative susceptibility 0.32, 95% CrI 0.21-0.48; relative infectivity 0.46, 95% CrI 0.29-0.70)(Figure 4).

**Figure 4.**
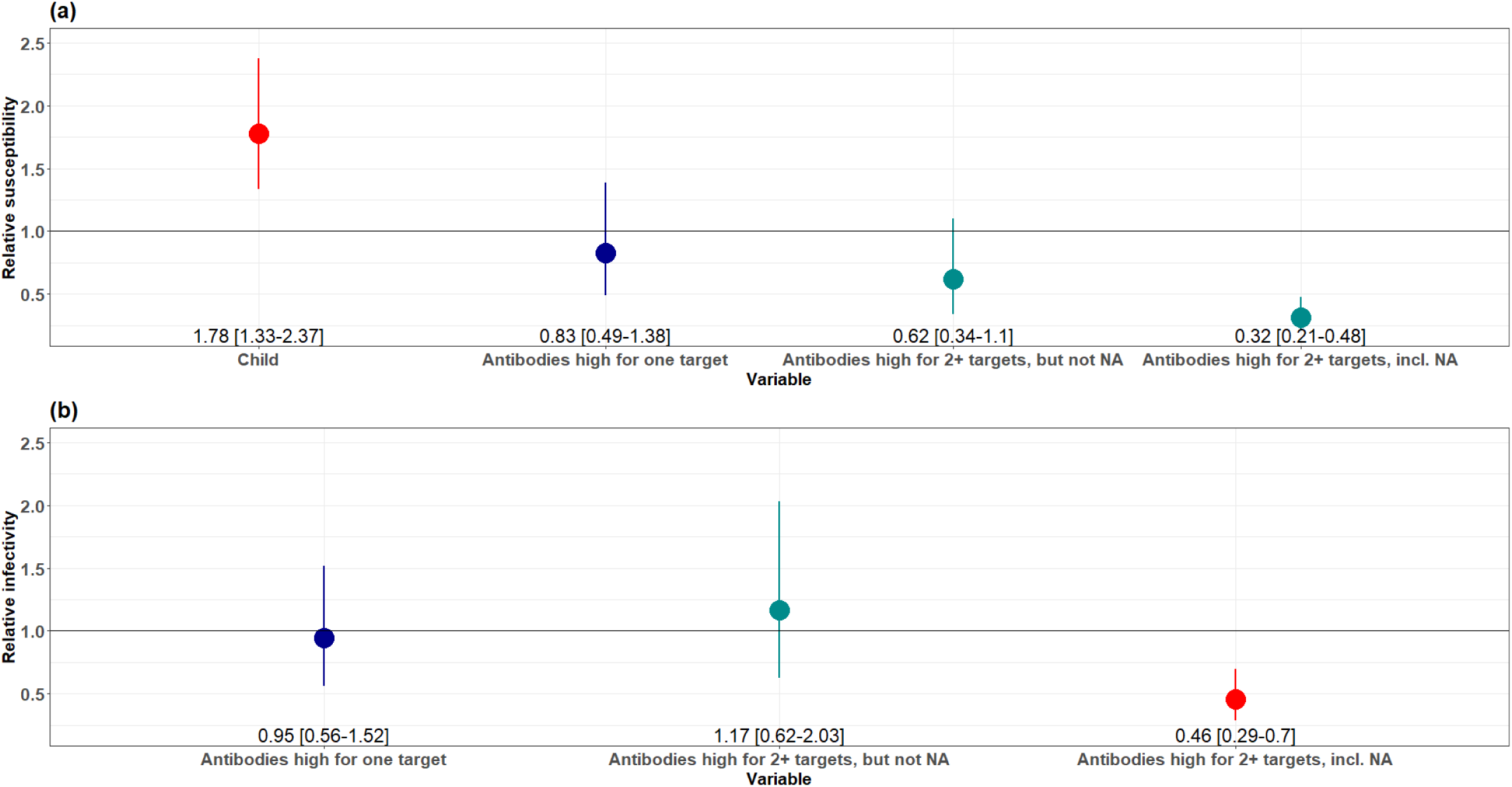
Effect of cumulative antibody levels on susceptibility and infectivity. Point estimate and 95% credible interval for (a) relative susceptibility to A/H3N2 as a function of age and categorical higher-versus-low antibody levels for zero (reference), one, two or more (not including NA), or two or more (including NA) antibody targets, and (b) relative infectivity of A/H3N2 as a function of categorical, higher-versus-low antibody levels for zero (reference), one, two or more (not including NA), or two or more (including NA) antibody targets.

### Effect of anti-NA antibodies in individuals with existing anti-HA antibodies

Finally, we asked whether the benefit of anti-NA immunity was only seen in those with little-to-no anti-HA immunity, or whether anti-NA immunity was beneficial even in individuals with high pre-existing antibody levels to HA. To do this, we investigated the relative contribution of anti-NA antibody levels on influenza virus A/H3N2 infections among individuals who have existing high anti-HA antibodies to understand what, if any, benefit that high anti-NA antibodies have on susceptibility and infectivity among individuals who already have some anti-HA immunity. Among individuals with high anti-HA head and/or anti-HA stalk preexisting antibodies, those who also had higher anti-NA antibody levels had reduced susceptibility (relative susceptibility 0.43, 95% CrI 0.30-0.61) and infectivity (relative infectivity 0.42, 95% CrI 0.27-0.67) to influenza A/H3N2 virus infection, relative to those with low anti-NA antibody levels.

### Simulation analyses

Simulating epidemics in households from the model, we found that the transmission model was able to capture the observed patterns of secondary attack rates (SARs) by household size, even among large households (Supplemental Figure 1). In households of size 4, the most common household size in this study, the observed SAR was 0.18, and the estimated SAR across 100 simulations was 0.19 (95% CrI 0.14-0.26).

When we used our inference framework on data simulated from our model with known parameter values, parameter values were recovered consistently and with little directional bias. The simulation value fell within the 95% credible interval in 85%+ of simulations for all parameters, except for the NA infectivity parameter (70%) (Figure 5). In sensitivity analyses, we checked that the association between higher anti-NA antibody levels and reduced infectivity remains under different assumptions about distribution of the incubation and infectivity periods (Supplemental Table 1).

**Figure 5.**
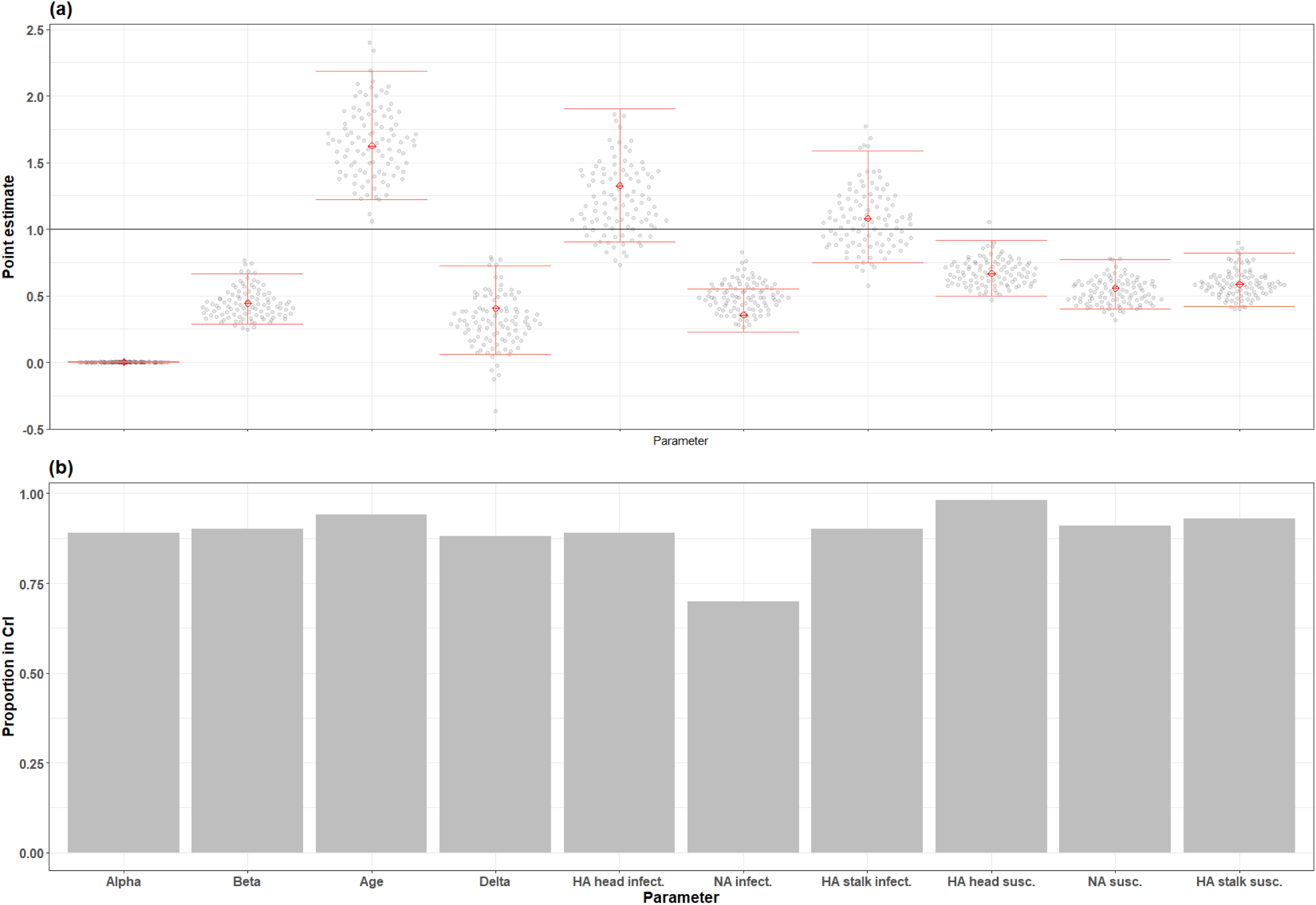
Parameter estimates in simulated datasets. (a) Point estimate and 95% credible interval for each parameter from the actual data, compared to the parameter point estimates recovered from running the MCMC on each simulated dataset. (b) proportion of simulations where the parameter point estimates recovered from the simulated dataset falls within the 95% credible interval of the parameters run on the actual data. Alpha is the community transmission parameter, beta is the household transmission parameter, and delta is a parameter for relating beta to household size.

## Discussion

Through intensive monitoring of households with known influenza A/H3N2 virus infection in combination with statistical transmission modeling, this study was able to reconstruct household transmission chains and assess the impact of individual-level factors, such as pre-existing antibody levels, on the susceptibility and infectivity of influenza A/H3N2 virus in a household setting.

Individuals who were infected with influenza A/H3N2 virus and who had high pre-existing antibodies against NA demonstrated reduced infectivity relative to infected individuals with low preexisting antibodies against NA. The magnitude of this difference is substantial, with high-anti-NA individuals having 64% (95% CrI 40-79%) reduced infectivity compared to low-anti-NA individuals. Importantly, preexisting antibodies against the HA head and HA stalk were not associated with reduced infectivity, indicating that a reduction in influenza infectivity may depend on anti-NA responses alone. This specificity is biologically plausible, as NA, not HA, is responsible for viral egress, and thus it is reasonable that anti-NA responses alone contribute to a reduction in overall viral load, viral shedding, and subsequent transmission^16^.

We found that all the factors of interest included in the transmission model, namely age and pre-existing antibody levels against the HA head, HA stalk, and NA, were associated with influenza susceptibility. Specifically, children aged 14 years and younger were 63% more susceptible to infection than adults aged 15 years and older. This is in line with a large body of work indicating that children are more susceptible to influenza virus infection but suggests that this susceptibility is not entirely due to a lack of influenza exposure history and immune response, as we observe a large association between age and susceptibility even when accounting for anti-HA head, anti-HA stalk, and anti-NA antibody levels^22,24–28^. Individuals with high preexisting antibodies demonstrated reduced susceptibility to influenza A/H3N2 virus infection, with reductions of 33%, 41%, and 44%, respectively, for antibodies against the HA head, HA stalk, and NA, which is in line with previous work on the correlates of protection against influenza viruses, including influenza A/H3N2^12,13,29,30^.

We found that susceptibility to and the infectivity of influenza A/H3N2 virus was not reduced when individuals had higher antibodies for only one target (HA head, HA stalk, or NA), nor were they reduced when individuals had higher antibodies for multiple HA targets but not NA. However, having higher antibodies for two or more targets, with one being NA, was associated with a 68% (52%-79%) reduction in susceptibility and 54% (30%-71%) reduction in infectivity, respectively, compared to individuals with low-to-undetectable antibodies against all targets. These findings emphasize the importance of generating robust, multi-epitope immune responses in vaccine-development efforts to generate vaccinations that protect adequately against infection and transmission. They also suggest that the induction of robust anti-NA immunity may be especially important.

Among individuals with existing immunity to HA, high anti-NA antibody levels are associated with a reduction in influenza A/H3N2 virus infectivity and susceptibility, indicating that the induction of better anti-NA response may be beneficial even in individuals with strong anti-HA immunity. Though current-generation influenza vaccines typically include a neuraminidase component, the immunogenicity of the NA component of vaccines is inconsistent, and anti-NA responses are often not utilized as an endpoint in the projection of vaccine efficacy against seasonal influenza^5,31^. These results suggest that, to generate next-generation influenza vaccines that are effective at reducing susceptibility as well as infectivity, the anti-NA response generated by vaccine candidates needs to be emphasized, measured, and assessed.

The secondary attack rate (the proportion of household contacts that become infected) in the study population was 22.3%, which is consistent with that found from other studies, especially given that there is a large proportion of children who have been associated with higher influenza SARs, in this population relative to that of other studies^28,32^. As expected, individuals who remained negative for influenza A/H3N2 virus throughout the monitoring period had higher initial levels for antibodies against the HA head, HA stalk, and NA when compared to individuals who became positive. Index cases had a similar distribution of pre-existing antibody levels compared to secondary cases for anti-HA head and stalk antibodies, indicating that there is likely not a large bias in the ascertainment of index cases vs. secondary cases relative to preexisting antibody levels. The small difference in distribution observed for NA may be due to differing age and symptom distributions between index cases and secondary cases.

This study is strengthened by the relatively large sample size, robust immunologic characterization of participants before and after infection, and methods that allow for probabilistic chains of transmission to be reconstructed to assess individual-level risk factors for infectivity and susceptibility, rather than assuming all secondary cases arise from the index case. Furthermore, the unvaccinated nature of the population allows us to specifically examine infection-induced immunity.

This study is limited by the low number of vaccinated participants, which makes stratification by vaccination status impossible; the relative contribution of anti-NA antibodies on A/H3N2 transmission may differ between vaccine-induced and infection-induced immunity in ways that we cannot assess. Furthermore, even though our modelling accounts for the possibility of community infections, we cannot rule out the possibility that some household contacts infected in the community might have been misattributed to in-household transmission; however, we would not expect this to be specific to higher or low-to-undetectable antibody levels for any of the targets and would not expect a directional bias in these estimates.

## Conclusions

Using data from two large household transmission studies, we found that, though pre-existing anti-HA head, stalk, and anti-NA antibodies are important for reduced susceptibility to influenza A/H3N2 virus infection, only anti-NA antibodies are associated with reduced infectivity in a household transmission setting. Our results suggest that the induction of a better humoral immune response against NA may improve next-generation vaccines’ effectiveness at preventing infection and disease and may reduce individual infectivity even in the event of a breakthrough infection. These findings reinforce the need for continued development of influenza vaccinations that target NA in addition to HA in order to develop next-generation influenza vaccines that protect against influenza virus infection and reduce influenza infectivity.

## Online Methods

### Study Population and Design

This study uses data from two household influenza transmission studies based in Managua, Nicaragua: the Household Influenza Transmission Study (HITS) and the Household Influenza Cohort Study (HICS). HITS is a case-ascertained study, meaning that influenza-positive individuals are identified, and other members of their household recruited for enrollment, that ran from 2012 to 2017, and HICS is a prospective household-based cohort study that began in 2017 and is currently ongoing. In both studies, influenza A/H3N2 virus-positive individuals, the index cases, are initially detected at a health center, where household members are enrolled (HITS) or activated (HICS) into intensive monitoring for a period of ∼14 days. During this period, household members are tested repeatedly for influenza virus, allowing for a reconstruction of likely transmission chains within each household. Blood samples are collected both at the beginning of the monitoring period and 30-45 days after^33^. These studies were approved by the institutional review boards at the Nicaraguan Ministry of Health and the University of Michigan and are in accordance with the Helsinki Declaration of the World Medical Association. Written consent to participate or parental permission was obtained for all participants; in children older than 6 years, verbal assent was obtained.

### Laboratory Methods

Nasal/oropharyngeal swabs collected from household members were tested for influenza virus with real-time reverse-transcription polymerase chain reaction (RT-PCR) using validated Centers for Disease Control and Prevention (CDC) protocols. If positive for influenza virus, subtype or lineage determination was performed using additional RT-PCR assays^34–36^. Several serological assays were conducted on each blood sample to measure the initial and final antibody levels against various influenza antigens; hemagglutination inhibition assays (HAIs), and enzyme-linked immunosorbent assays (ELISAs) against full-length HA, the HA stalk, and NA. Details about the specific antigens used for each assay are available in the supplement (Supplemental Table 2).

### Statistical Methods

Preexisting antibody levels were divided into quartiles, with the lowest quartile corresponding to low antibody levels and the remaining quartiles corresponding to high antibody levels. The distribution of initial antibody levels for index cases, secondary infections within the household of the index case, and uninfected household members were compared using two-sided Wilcoxon rank-sum tests.

We used a mathematical model to assess the impact of individual-level age and immune characteristics and contact structure on the person-to-person probability of transmission. The model estimated the risk of transmission between all household members including the risk from secondary cases. The risk of transmission from an infected individual depended on time after infection with by lognormal distribution^23,37,38^. This risk was modulated by infectivity factors, namely individual pre-existing anti-HA head, anti-HA stalk, and anti-NA antibody levels of the infector. It was also modulated by the susceptibility factors being the individual characteristics of the susceptible contact, namely individual age and pre-existing anti-HA head, anti-HA stalk, and anti-NA levels. Finaly the risk depended on the household size. We also estimated the risk of infection from the community. This transmission model accounts for chains of transmission within households, namely that additional household infections beyond the index case may occur due to community transmission or due to infection from a non-index household member, which is advantageous relative to other common statistical approaches to transmission, approaches including logistic models^23,24,39,40^. Additional information about the transmission model, including the functional forms and mathematical formalism, are included in the Supplement.

Model parameters were estimated using Bayesian Markov chain Monte Carlo (MCMC)^23,41^. The statistical model has a hierarchical structure with three levels^23^: i) the observation level ensures consistency between observed and augmented data (based on the probabilistic distribution assumed for the incubation period^23,41–43^), ii) the transmission model (described above), characterizes within household transmission dynamics, iii) the prior model describe prior distribution for model parameters.

Transmission parameters and augmented times of infection were iteratively updated using a Metropolis-Hastings algorithm^23,41–45^. Each MCMC chain was iterated 50,000 times, and the first 500 iterations were burned out. We report the median of the posterior distribution with the 95 credible interval (Crl) for each estimated transmission parameter.

Additionally, 100 datasets of simulated infection events were generated by an agent-based model by retaining household structure, index case assignment, and individual characteristics such as age and antibody levels. Infection events at each time step were drawn randomly from the probability of transmission derived from the transmission model. Transmission parameters used were the posterior medians. We assessed model adequacy by comparing secondary attack rates at each household size generated by the model to those generated by the observed data, and examined the bias of estimated parameters, including the proportion of the 95% credible interval from the original posterior distributions that covered the median simulation values. Analyses were conducted using SAS 9.4, R version 4.3.1-4.3.2, and Visual Studio Code version 1.87.2

## Acknowledgements

This work was supported by the National Institutes of Allergy and Infectious Diseases through the Collaborative Influenza Vaccine Innovation Centers [75N93019C00051 to F.K. and A.G.]the St. Jude Center of Influenza Research and Surveillance [HHSN272201400006C to A.G.]; the St. Jude Center of Excellence for Influenza Research and Response [75N93021C00016 to A.G.]and grant R01 AI120997 to A.G. Aubree Gordon is supported by the Biosciences Initiative at the University of Michigan through a Mid-career Biosciences Faculty Achievement Award (MBioFAR). S.C. acknowledges support by the European Commission under the EU4Health programme 2021-2027, Grant Agreement – Project: 101102733 — DURABLE, the Laboratoire d’Excellence Integrative Biology of Emerging Infectious Diseases program (grant ANR-10-LABX-62-IBEID) the European Union’s Horizon 2020 research and innovation programme under VEO grant agreement No. 874735, and the INCEPTION project (PIA/ANR16-CONV-0005). The funding agencies had no role in the design and conduct of the study, collection, management, analysis or interpretation of the data; preparation, review, or approval of the manuscript; or decision to submit the manuscript for publication.

We thank the study participants and the many dedicated study personnel in Nicaragua at the Centro Nacional de Diagnóstico y Referencia and the Sócrates Flores Vivas Health Center.

## Author Contributions

Gregory Hoy: Data curation; formal analysis; methodology; software; validation; visualization; writing—original draft; writing—review and editing. Thomas Cortier: formal analysis; methodology; software; validation; visualization; writing-original draft; writing-review and editing. Hannah E. Maier: data curation; methodology; software; validation; visualization; writing-review and editing. Guillermina Kuan: Investigation; project administration; writing—review and editing. Roger Lopez: Investigation; writing—review and editing. Nery Sanchez: Investigation; writing—review and editing. Sergio Ojeda: Investigation; project administration; writing—review and editing. Miguel Plazaola: Investigation; project administration; writing—review and editing. Daniel Stadlbauer: Data curation; writing—review and editing. Abigail Shotwell: data curation; writing-review and editing. Angel Balmaseda: Investigation; methodology; supervision; project administration; writing—review and editing. Florian Krammer: funding acquisition; investigation;; supervision; writing—review and editing. Simon Cauchemez: Conceptualization; investigation; methodology; project administration; resources; supervision; writing—review and editing. Aubree Gordon: Conceptualization; funding acquisition; investigation; methodology; project administration; resources; supervision; writing—review and editing.

## Conflict of interest statement

The Icahn School of Medicine at Mount Sinai has filed patent applications relating to SARS-CoV-2 serological assays, NDV-based SARS-CoV-2 vaccines influenza virus vaccines and influenza virus therapeutics which list Florian Krammer as co-inventor. Mount Sinai has spun out a company, Kantaro, to market serological tests for SARS-CoV-2 and another company, CastleVax, to develop SARS-CoV-2 vaccines. Florian Krammer is a co-founder and scientific advisory board member of CastleVax. Florian Krammer has consulted for Merck, Curevac, Seqirus, GSK and Pfizer and is currently consulting for 3rd Rock Ventures, Gritstone and Avimex. The Krammer laboratory is collaborating with Dynavax on influenza vaccine development. Aubree Gordon has served on an RSV vaccine advisory board for Janssen.

## Inclusion and ethics in global research statement

This study has included local researchers throughout the research process and is locally relevant as determined in partnership with local partners. Roles and responsibilities were agreed upon amongst collaborators ahead of the research, including capacity building for local researchers. This research was approved by the Nicaraguan Ministry of Health.

## Data availability statement

The data that support the findings of this study are available from the corresponding author upon reasonable request and following IRB approval.

## Code availability statement

Code supporting the findings of this research can be found at https://github.com/gehoy/H3N2_NA_transmission.

## Notes

### Author Declarations

This study was approved by the institutional review boards at the Nicaraguan Ministry of Health and the University of Michigan and is in accordance with the Helsinki Declaration of the World Medical Association. Written consent to participate or parental permission was obtained for all participants; in children older than 6 years, verbal assent was obtained.

