## Supplement for "Anti-Neuraminidase Antibodies Reduce the Susceptibility to and Infectivity of Influenza A/H3N2 Virus"

^1^School of Public Health, University of Michigan, Ann Arbor, Michigan, USA; ^2^Mathematical Modelling of Infectious Diseases Unit, Institut Pasteur, Université Paris Cité, UMR 2000 CNRS, Paris, France; ^3^Collège Doctoral, Sorbonne Université, Paris, France; ^4^Sustainable Sciences Institute, Managua, Nicaragua; ^5^Centro de Salud Sócrates Flores Vivas, Ministry of Health, Managua, Nicaragua; ^6^Laboratorio Nacional de Virología, Centro Nacional de Diagnóstico y Referencia, Ministry of Health, Managua, Nicaragua ^7^Department of Microbiology, Icahn School of Medicine at Mount Sinai, New York, New York, USA ^8^Center for Vaccine Research and Pandemic Preparedness (C-VaRPP), Icahn School of Medicine at Mount Sinai, New York, NY, USA. ^9^Department of Pathology, Molecular and Cell-Based Medicine, Icahn School of Medicine at Mount Sinai, New York, NY, USA. ^10^Ignaz Semmelweis Institute, Interuniversity Institute for Infection Research, Medical University of Vienna, Vienna, Austria

*Denotes co-senior authors

**Table of Contents**

**Supplemental Table 1. Sensitivity analyses for model assumptions**	**3**

**Supplemental Figure 1. Secondary attack rate in actual and simulated datasets**	**4**

**Supplemental Table 2. Antigens used for immunologic assays**	**5**

**Supplemental Methods**	**6**

**References**	**10**

*Supplemental Figure 1. Model diagnostics of primary model*

| **Parameter** | **Relative estimate for NA infectivity** |
| --- | --- |
| Default model | 0.36 (0.23-0.53) |
| Incubation 0.6 days (SD 0.8) | 0.36 (0.22-0.54) |
| Incubation 2 days (SD 1.6) | 0.38 (0.27-0.58) |
| Infectivity 2.5 days (SD 1.5) | 0.49 (0.32-0.70) |
| Infectivity 5 days (SD 2.5) | 0.24 (0.13-0.42) |

*Supplemental Table 1. Sensitivity analyses for model assumptions*

| **Parameter** | **Relative estimate for NA infectivity** |
| --- | --- |
| Default model | 0.36 (0.23-0.53) |
| Incubation 0.6 days (SD 0.8) | 0.36 (0.22-0.54) |
| Incubation 2 days (SD 1.6) | 0.38 (0.27-0.58) |
| Infectivity 2.5 days (SD 1.5) | 0.49 (0.32-0.70) |
| Infectivity 5 days (SD 2.5) | 0.24 (0.13-0.42) |

*Supplemental Figure 1. Secondary attack rate in actual and simulated datasets*
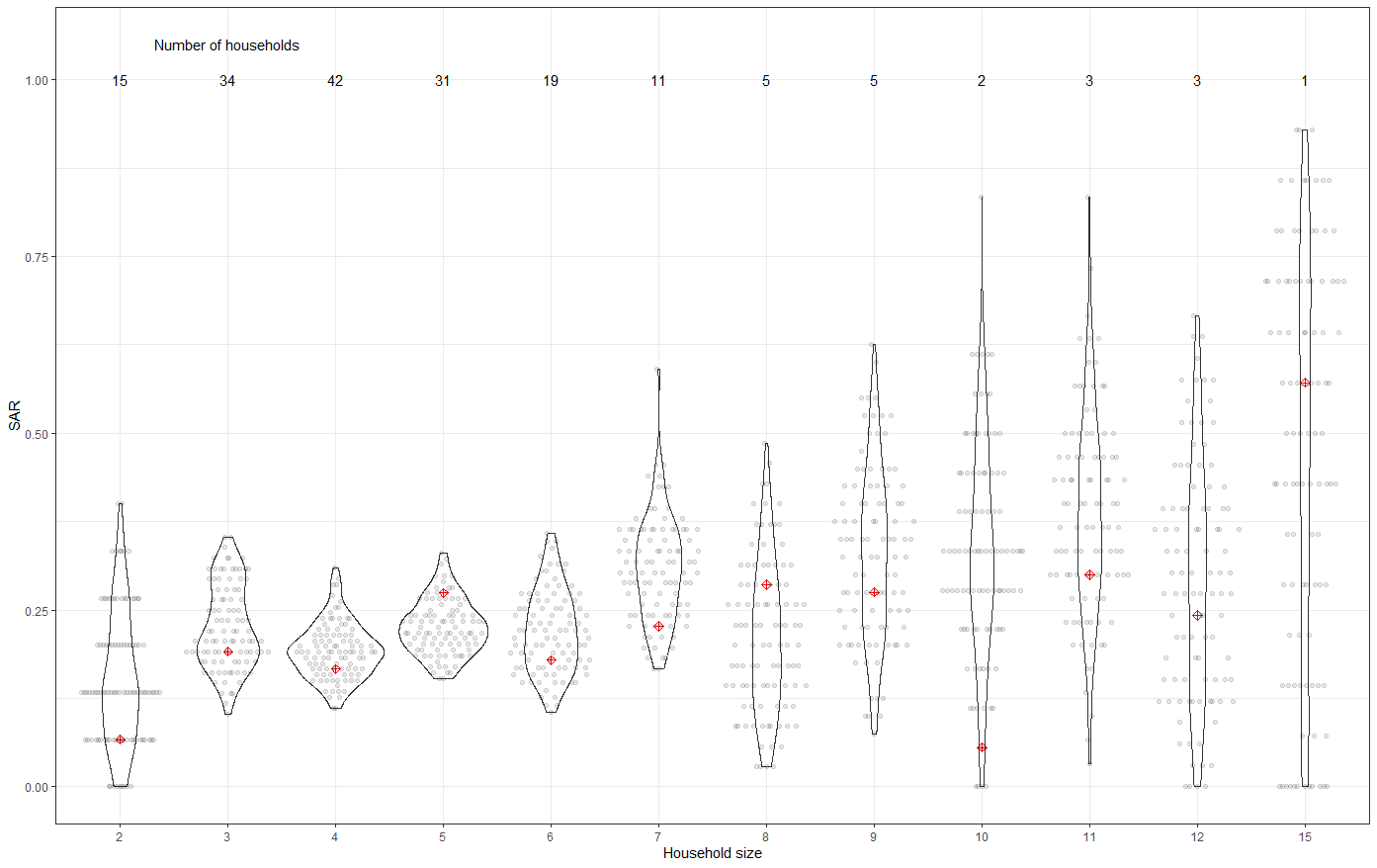

Secondary attack rates in actual and simulated datasets, by household size. The true secondary attack rate for the measured data is indicated by the red dots, and the calculated secondary attack rates from each simulated dataset is indicated by the grey dots; the violin plots indicate the distribution of secondary attack rates across simulated datasets.

*Supplemental Table 2 Antigens used for immunologic assays*

| **Assay** | **Antigen** |
| --- | --- |
| Hemagglutination inhibition assay | A/Hong Kong/4801/2014 |
| H3 ELISA | A/Hong Kong/4801/2014 |
| N2 ELISA | A/Hong Kong/4801/2014 |
| Stalk ELISA | cH7/3 with A/Anhui/1/2013 as the head domain and A/Hong Kong/4801/2014 as the stalk domain |

**Supplemental Methods**

**Data considered in the transmission model**

Denote *i* the index of an individual in a household, their age $a_{i}$, their pre-existing anti-HA-head antibody level $hha_{i}$, their pre-existing anti-HA-stalk antibody level $sha_{i}$, their pre-existing anti-NA antibody level $na_{i}$, their infection status $C_{i}$(1 if infected, 0 otherwise), their symptom onset date $S_{i}$,$T_{i}$ their unobserved (and augmented) infection date. Denote the end of the follow-up period $t_{end}$.

**The transmission model**

**The risk of transmission between two household members**

With one infected individual *i* and one susceptible contact *j* in a household of size *n*, the instantaneous risk that case *i* infects susceptible contact *j* at time t is:

${\lambda_{{i\to}_{.}j}\left( t \right)=\beta\left( \frac{5}{n} \right)^{\delta}\mu_{a}(a_{j})\mu_{hha}(hha_{j})\mu}_{sha}(sha_{j})I_{hha}\left( hha_{i} \right)I_{sha}\left( sha_{i} \right)I_{na}\left( na_{i} \right)f(t-T_{i})$

where

- $\beta$ is the scaling parameter of the household risk of infection
- $n^{\delta}$ is the household size dependency of the transmission risk, where $\left( \frac{5}{n} \right)^{\delta}$refers to the average household size of 5
- $\mu_{a}(a_{j})$ is the relative transmission risk related to age, considering to groups (children: <15 years old; adults: 15 years and older). The reference is the adult group.
- $\mu_{hha}(hha_{j})$ is the relative susceptibility related to pre-existing anti-HA head antibodies, considering to groups (low-to-undetectable: HAI titers of 1:5 or less; higher: HAI titers greater than 1:5). The reference is individuals with low-to-undetectable HAI titers.
- $\mu_{sha}({sha}_{j})$ is the relative susceptibility related to pre-existing anti-HA stalk antibodies, considering to groups (low-to-undetectable: HA stalk AUCs of 15.3 or less; higher: HA stalk AUCs of greater than 15.3). The reference is individuals with low-to-undetectable HA stalk antibodies.
- $\mu_{na}({na}_{j})$ is the relative susceptibility related to pre-existing anti-NA antibodies, considering to groups (low-to-undetectable: NA AUCs of 21.8 or less; higher: anti-NA AUCs of greater than 21.8: . The reference is individuals with low-to-undetectable NA antibodies
- $I_{hha}\left( hha_{j} \right)$ is the relative infectivity related to pre-existing anti-HA head antibodies considering to groups (low-to-undetectable: HAI titers of 1:5 or less; higher: HAI titers greater than 1:5). The reference is individuals with low-to-undetectable HAI titers $I_{sha}({sha}_{j})$ is the relative infectivity related to pre-existing anti-HA stalk antibodies, considering to groups (low-to-undetectable: HA stalk AUCs of 15.3 or less; higher: HA stalk AUCs of greater than 15.3).. The reference is individuals with an anti-HA stalk ELISA AUC of at the 25^th^ percentile (15.3) or less.
- $I_{na}({na}_{j})$ is the relative infectivity related to pre-existing anti-HA stalk antibodies considering to groups (low-to-undetectable: NA AUCs of 21.8 or less; higher: anti-NA AUCs of greater than 21.8:. The reference is individuals with an anti-HA stalk ELISA AUC of at the 25^th^ percentile (21.8) or less. The reference is individuals with low-to-undetectable NA antibodies
- $f(t-T)$ is the generation time (i.e. the time between infector infection time $T_{i}$ to secondary transmission time $T_{j}$) density. This was assumed to follow a gamma distribution with a mean duration of 4 days (SD 2 days)^1–3^.

**Risk of infection for contact j**

The total risk of infection for individual *j* in household *k* at time *t* is dependent on the overall force of infection from all infected cases in the household, along with the risk of community transmission α that we assume to be constant over time.

$$\lambda_{j}\left( t \right)=\mu_{a}(a_{j})\mu_{hha}(hha_{j})\mu_{sha}({sha}_{j})\alpha+\sum_{i\in\{T_{i}<t\}} \lambda_{{i\to}_{.}j}\left( t \right)$$

The probability of transmission from the community for period of T days is equal to

$Pcommunity(T) =1-exp(-$ $\alpha$ $T)$

**The likelihood of the transmission process in household k**

We denote $Sus_{k}$ the group of non infected individuals ( $C_{i}$=0), $Inf_{k}$the group of infected individuals ($C_{i}$=1) and assume i=1 for the index case of the household. $\psi$ is the vector of model parameters.

Conditional on the first infection time $T_{1}$, given symptom onset times *S*, infection events $T$and infection status $C$, the likelihood of the transmission process is :

$$P(S,C,T|\psi)=\prod_{i\in{Sus}_{k}} P(C_{i}=0|\psi)\prod_{i\in{Inf}_{k}-1} P(T_{i},C_{i}=1|\psi)\prod_{i\in{Inf}_{k}} P(T_{i}|S_{i})$$

#### The contribution to the likelihood of a uninfected individuals

This is the probability for an individual to avoid infection for the all follow up period.

$P(C_{i}=0|\psi)$=$exp(-\int_{T_{1}}^{t_{end}} \lambda_{i}(t)dt)$

#### The likelihood of infected contact

This is the probability of escaping infection up to $T_{i}$ and then becoming infected at $T_{i}$

$P(C_{i}=1,T_{i}|\psi)$=$\lambda_{i}(T_{i})exp(-\int_{T_{1}}^{T_{i}} \lambda_{i}(t)dt)$

#### The likelihood of symptom onset time given infection time

For all infected cases, the likelihood of symptom onset time $S_{i}$ given infection time $T_{i}$ is

$P(S_{i}|T_{i})=$g($S_{i}-T_{i}$)

where the incubation period follows a gamma distribution g with a mean incubation of 1.0 day (SD 1.2 days)^4–6^

**Dealing with missing infection times**

The transmission process and its likelihood depends on unobserved infection events. We augmented these unobserved infection events $T_{i}$ from observed symptom dates $S_{i}$.

**MCMC algorithm**

We utilized a Bayesian framework with MCMC augmentation to explore the posterior distributions of transmission parameters and augmented infection times. Transmission parameters were updated using a Markov chain Monte Carlo algorithm with the Metropolis-Hastings algorithm, with log-normal proposals for positive parameters and normal proposals for others. Proposals standard deviations were tuned to ensure that the acceptance rate was between 20% and 40% for all parameters. Infection times were augmented for each iteration of the MCMC chain. The proposal used to update the incubation period of an individual was a gamma distribution with mean 1.0 days and standard deviation 1.2 days^4–6^.

**Prior distributions of transmission parameters**

We use a uniform prior between 0 and 1 for the risk from the community $\alpha$. A uniform prior between 0 and 5 for $\beta$ and a uniform prior between -3 and 3 for $\delta$ $.$ Prior distributions for all susceptibility and infectivity factors were assumed to be lognormal with standard deviations of 1.0.

**Programming**

The MCMC algorithm was coded in C++ using Visual Studio Code version 1.89.1. Chains were run for 50,000 iterations and recorded every 20 iterations. Posterior parameter distributions were sampled from the MCMC after discarding a burn-in of 500 iterations. Acceptance rate and convergence was assessed visually.
